# Outcomes of Ivermectin in the treatment of COVID-19: a systematic review and meta-analysis

**DOI:** 10.1101/2021.01.26.21250420

**Authors:** Alex Castañeda-Sabogal, Diego Chambergo-Michilot, Carlos J. Toro-Huamanchumo, Christian Silva-Rengifo, José Gonzales-Zamora, Joshuan J. Barboza

## Abstract

**Background:** To assess the outcomes of ivermectin in ambulatory and hospitalized patients with COVID-19.

**Methods:** Five databases and websites for preprints were searched until January 2021 for randomized controlled trials (RCTs) and retrospective cohorts assessing ivermectin versus control in ambulatory and hospitalized participants. The primary outcome was overall mortality. Secondary outcome was recovered patients. For meta-analysis, random-effects and inverse variance meta-analyses with logarithmic transformation were performed. ROBINS-I for cohort studies, and the Cochrane Risk of Bias 2.0 tool for trials were used. The strength of evidence was assessed using GRADE.

**Results:** After the selection, twelve studies (five retrospective cohort studies, six randomized clinical trials and one case series), were included. In total, 7412 participants were reported, the mean age was 47.5 (SD 9.5) years, and 4283 (58%) were male. Ivermectin was not associated with reduced mortality (logRR: 0.89, 95% CI 0.09 to 1.70, p = 0.04, I^2^= 84.7%), or reduced patient recovery (logRR 5.52, 95% CI -24.36 to 35.4, p = 0.51, I^2^ = 92.6%). All studies had a high risk of bias, and showed a very low certainty of the evidence.

**Conclusions:** There insufficient certainty and quality of evidence to recommend the use of ivermectin to prevent or treat ambulatory or hospitalized patients with COVID-19.

## INTRODUCTION

Since the first reported case of severe respiratory syndrome coronavirus 2 (SARS-CoV-2), in December 2019 in Wuhan, China; cases of coronavirus disease (COVID-19) have increased exponentially, with more than 95 million infected people worldwide until January 18, 2021 (https://coronavirus.jhu.edu/map.html). This high volume of COVID-19 cases has led to several problems including an overburdened health system, and a worrisome shortage of healthcare personnel. In this setting, finding an effective therapy against SARS-CoV-2 has become an urgent need (1)

The current treatment of COVID-19 has been limited to general supportive care, because studies evaluating the efficacy of treatment in patients with COVID-19 have had several limitations and no treatment has demonstrated strong evidence for widespread recommendation (2).

Ivermectin is a semisynthetic anthelmintic agent that selectively binds to glutamate-gated chloride ion channels found in nerve and muscle cells of invertebrates (3). However, an antiviral activity in RNA and DNA viruses has also been reported (4).

Caly, et al. conducted an in vitro study, in which they inoculated the SARS-CoV-2 virus in Vero/hSLAM cells, and found that Ivermectin at a dose of 4 µM reduced the viral load after 48 hours. This finding encouraged the conduction of studies aimed to evaluate the clinical effectiveness of Ivermectin (5).

On the other hand, Virginia D. Schmith et al. developed a pharmacokinetic model, with transit absorption, first-order elimination and weight as covariates in the central volume of distribution and clearance. These authors used approved doses of 200 µg/kg (in 3 mg increments); 120 mg MD and 60 mg three times weekly (every 72 hours) and concluded that the inhibitory concentration (IC_50_) of ivermectin was not expected to inhibit SARS-CoV-2 in lung tissue even simulating with 10 times the approved dose in humans. Similar findings were reported by Caly et al (5, 6).

We conducted a systematic review and meta-analyses to evaluate the clinical efficacy of ivermectin for the treatment of COVID-19. This will provide clinicians with an overview of the scientific evidence on a potential treatment option, which will help in the clinical management of COVID-19 patients.

## METHODS

### Protocol

This systematic review was reported according to the Preferred Reporting Items for Systematic Reviews and Meta-Analyses (PRISMA) (7).

### Data sources

We searched PubMed, Scopus, Web of Science, Ovid-Medline, Embase, websites for preprints/preproofs (“Other sources” ; https://www.medrxiv.org, https://preprints.scielo.org/index.php/scielo, https://www.biorxiv.org, https://arxiv.org), websites for protocols of clinical trials (https://clinicaltrials.gov). We performed a search strategy for each database. The complete search strategy is found in *Supplementary file*. We included all the original published studies (either as preprints or in scientific journals) of clinical trials, non-randomized studies of intervention, and retrospective cohorts, without language restrictions, from inception to January 21, 2020; that have included patients (ambulatory or hospitalized) with COVID-19, and have compared a group that received ivermectin with a group that did not; regardless of their study design. Systematic reviews, narrative reviews, conference proceedings, editorials, and letters to the editor without original data were excluded.

### Outcomes

Primary outcome was overall mortality. Secondary outcome was recovered patients.

### Study selection

Two authors (JJB, DCM) independently screened search results by title and abstract according to the inclusion and exclusion criteria, using a web program *Rayyan* (rayyan-qcri.org). Also, two authors (JJB, DCM) independently assessed relevant studies and selected by full-text for the next phase of assessment. Discrepancies were consulted with another author (ACS), and a consensus was reached. The selection of articles in each stage of the review process was made using the Endnote X9 software.

### Data extraction

Two authors (JJB, DCM) independently extracted the data using pre-piloted Excel spreadsheets. Again, discrepancies were consulted with another author (ACS). The data extracted from each study were: Author, year, country, type of study, number of patients with ivermectin treatment, treatment/comparison or control arm, characteristics and condition of the patient when the treatment was received, methods of assessment and confounding variables, outcomes, and absolute effect of ivermectin versus control. Regarding the outcome of recovered patients, this variable was evaluated according to the criteria considered by the authors of each included study.

### Risk of bias assessment

Two investigators (JJB, DCM) independently assessed the risk of bias by using the ROBINS-I (Risk Of Bias In Non-Randomized Studies of Interventions) tool (8) for cohort studies and the Cochrane Risk of Bias 2.0 tool (9) for trials; disagreements were resolved by discussion with a third investigator (ACS).

### Data synthesis and statistical analysis

We assessed the certainty of evidence using the GRADE methodology (10). When possible, we meta-analyzed results of RCTs and non-randomized studies that have used methods to control by possible confounders.

Random-effects models with Hartung-Knapp adjustment for random effects model, and the inverse variance method were used for all meta-analyses. Effects of ivermectin were described with log relative risks (LogRRs) with 95% confidence intervals (Log RR 95% CIs) for dichotomous outcomes in the observational studies that were assessed. Heterogeneity among studies was assessed using the I^2^ statistic: 0–30% meant low, 30– 60% moderate, and >60% high heterogeneity (11). Subgroup analysis by year (<60 years vs >60 years), condition (outpatient vs inpatient), and need of oxygen support (with vs without oxygen support) was proposed; however, due to the insufficient published evidence, these aspects were not evaluated. A sensitivity analysis was performed excluding those studies without adjustment for confounding. (12).

### Ethical considerations

This is a systematic review of published and open information, in which no human subjects participated. No ethics committee approval was required.

## Results

### Selection of studies

The search yielded 532 results. After duplicates were excluded, 232 titles and abstracts were reviewed, 210 of these were excluded, and 22 scientific papers were evaluated in detail.

Finally, 12 studies were included in the qualitative synthesis (13-24), and five studies were included in the quantitative synthesis (Figure 1). Most studies were pre-print studies and two studies (18, 22) were anticipated results of clinical trials protocols.

**Figure 1.**
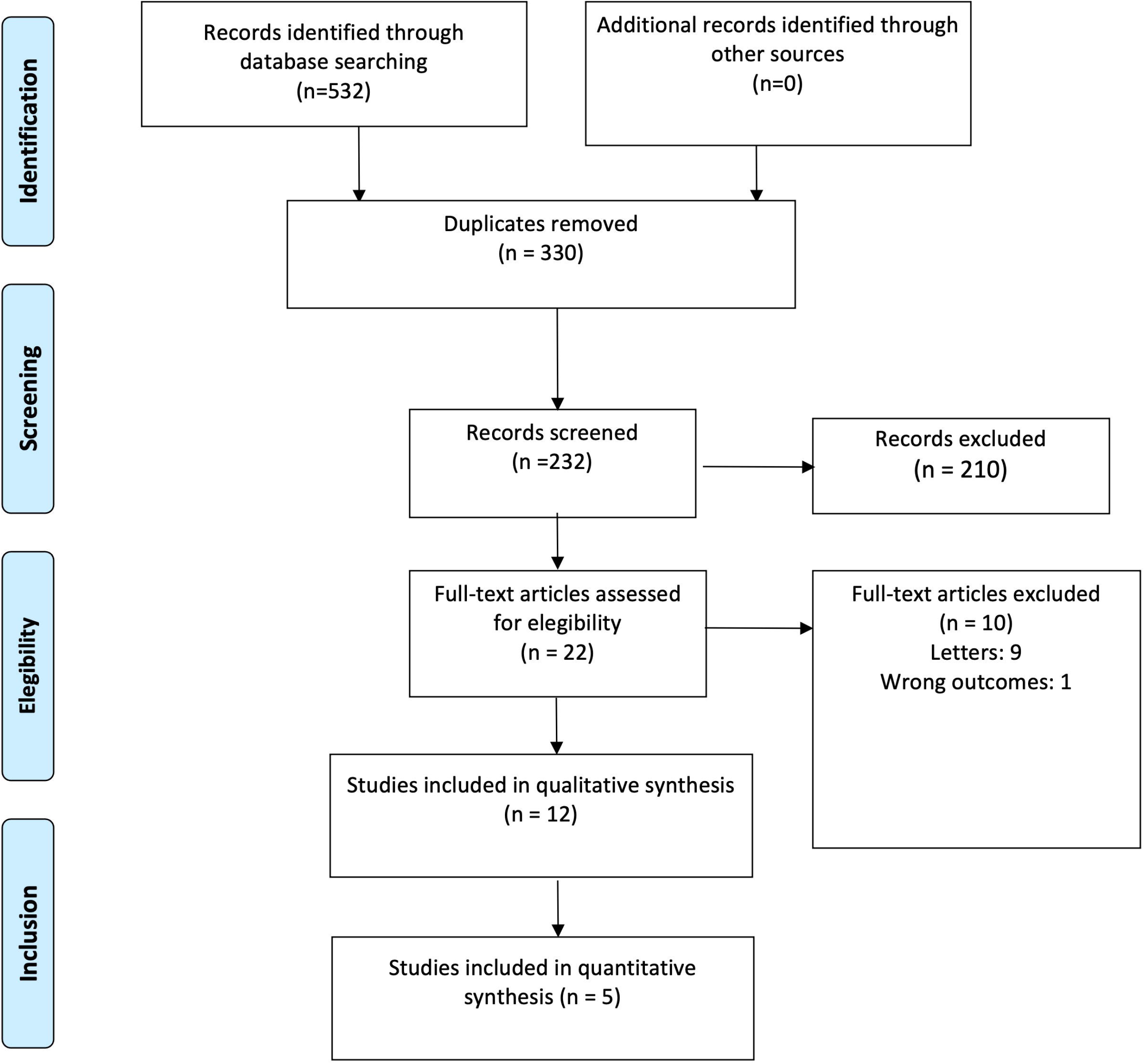
PRISMA flowchart for included studies.

### Characteristics of the studies included

Main characteristics of included studies are summarized in Table 1. Two studies were located in USA (21, 22), two from South America (15, 23), one from Iraq (16), two from Spain, one from Iran, and four from Bangladesh (13, 17, 18, 20). Five retrospective cohort studies (14, 16, 17, 21, 23, 25), six clinical trials (13, 18-20, 22, 24), and one case series (15) were found. There were 7412 reported participants, the mean age was 47.5 (SD 9.1) years, and 4283 (58%) were male. The treatment was ivermectin (alone or with azithromycin, hydroxychloroquine, dexamethasone, enoxaparin, aspirin or dicloxacillin). Only one study reported no control group (15). Most patients were hospitalized and confirmed COVID-19 by RT-PCR, except for individuals included in one study evaluating asymptomatic families (22). Regarding methods of assessment and confounding variables, two studies analyzed the confounding variables by propensity score weighting and adjusted by age, sex, location, type of admission, comorbidities, antibiotics applied and other drugs (21, 23). One study assessed these variables by logistic and Cox regression adjusted by age, sex, comorbidities and use of other drugs (21). One study assessed the variables by Kaplan Meier survival curve, adjusted by age, gender, and severity (16). Finally, eight studies were not adjusted by confounding variables because they only applied bivariate analysis (13-15, 17-20, 22). Different outcomes were evaluated by included studies, and most studies have assessed mortality and recovered patients as the primary outcome. Only four studies did not describe data regarding mortality and recovery in their analysis (14, 15, 22, 26).

**Table 1.**
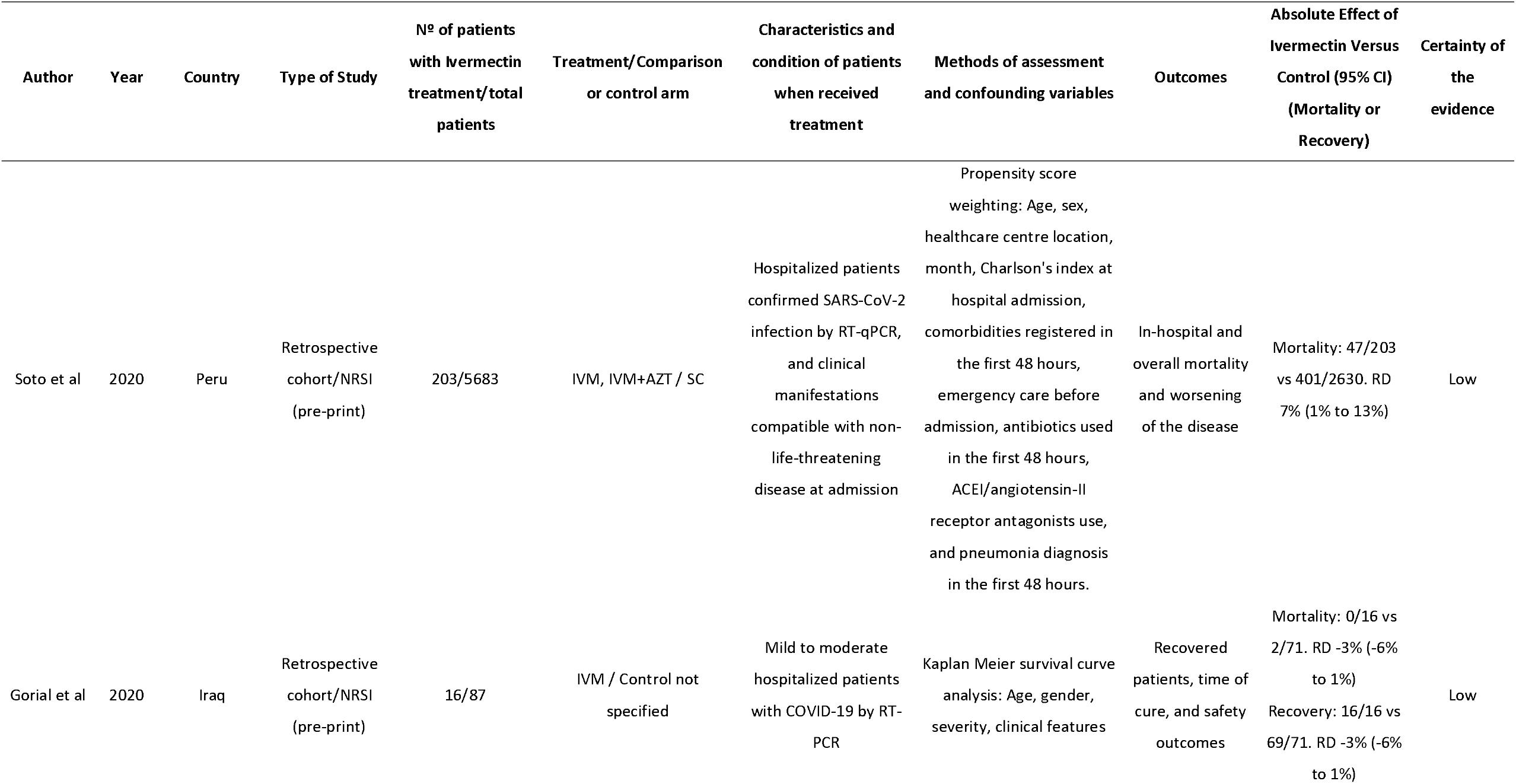
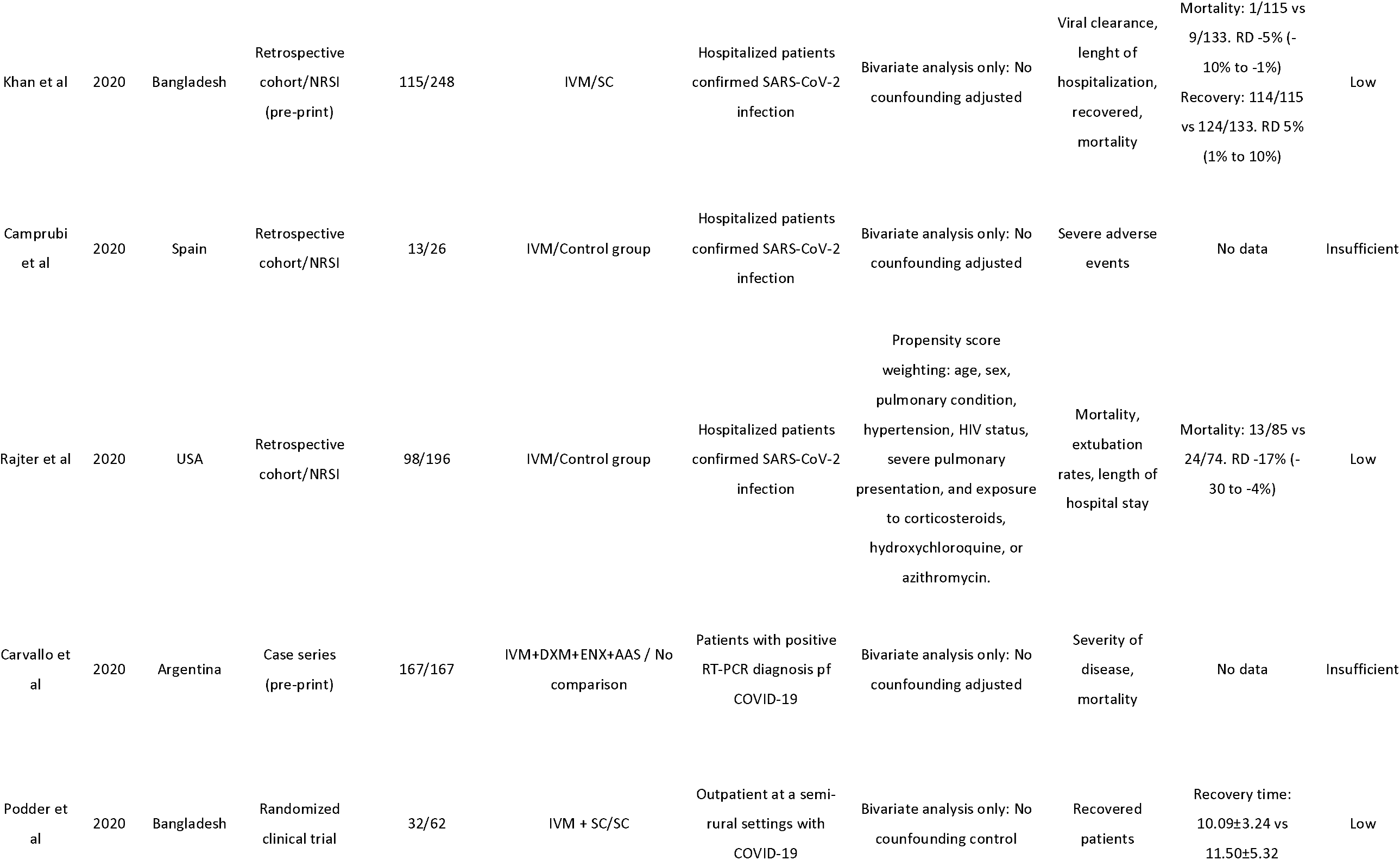
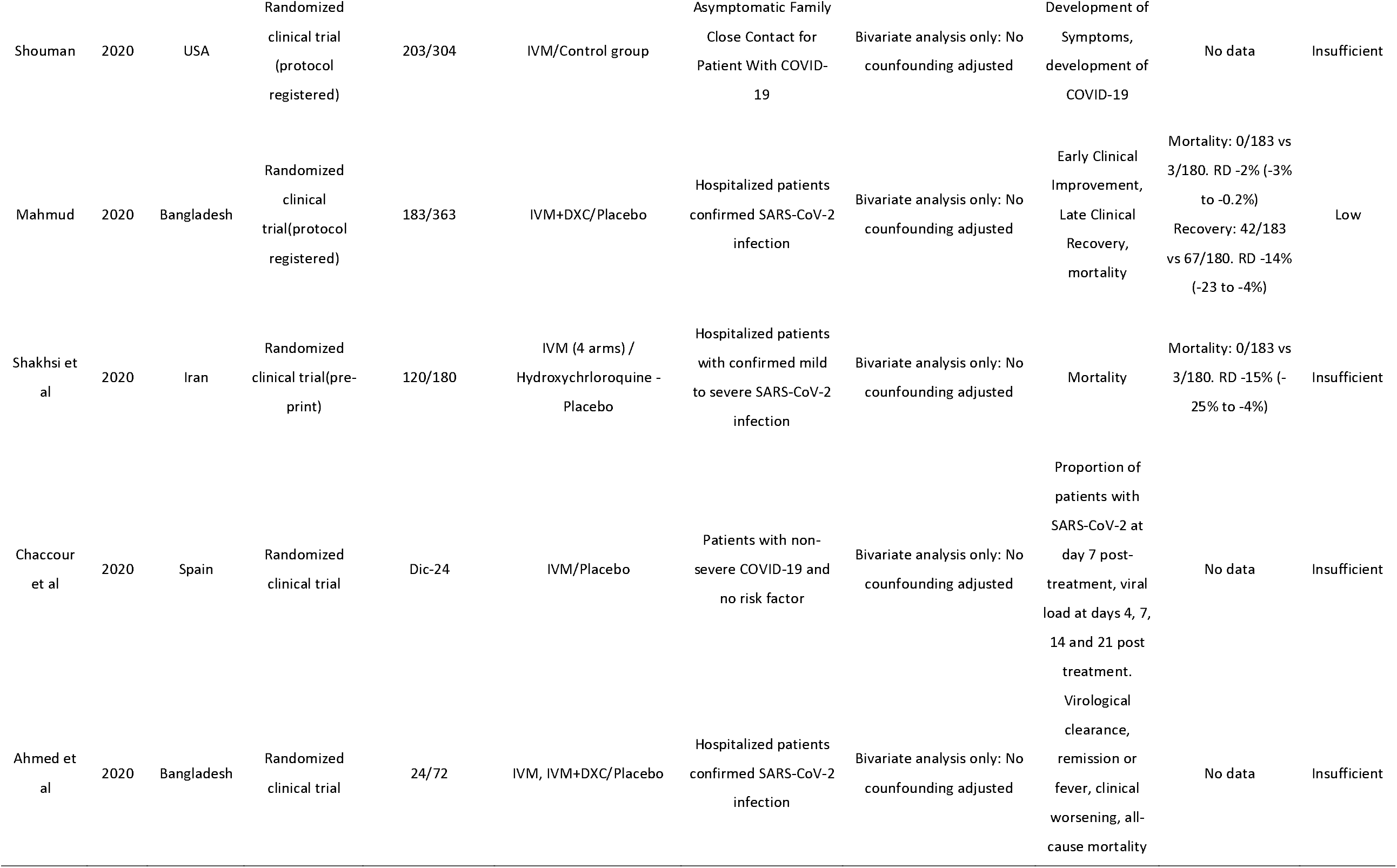
Characteristics of studies included.

### Assessment of risk of bias

Five RCT had high risk of bias due to missing outcome data (18-20, 22, 26). Four cohorts had serious risk of bias: Two studies were at serious risk of bias due to classification of interventions (16, 23), and two studies had critical risk of bias due to confounding (14, 17).

### Primary and secondary outcomes of ivermectin in patients with COVID-19

In this analysis with four pre-print retrospective studies, and high risk of bias, ivermectin is not associated with reduced mortality (logRR 0.89, 95% CI 0.09 to 1.70, p = 0.04, I^2^= 84.7%, Figure 2a). Additionally, ivermectin was not associated with reduced patient recovery (logRR 5.52, 95% CI -24.36 to 35.4, p = 0.51, I^2^ = 92.6%, Figure 2b).

**Figure 2.**
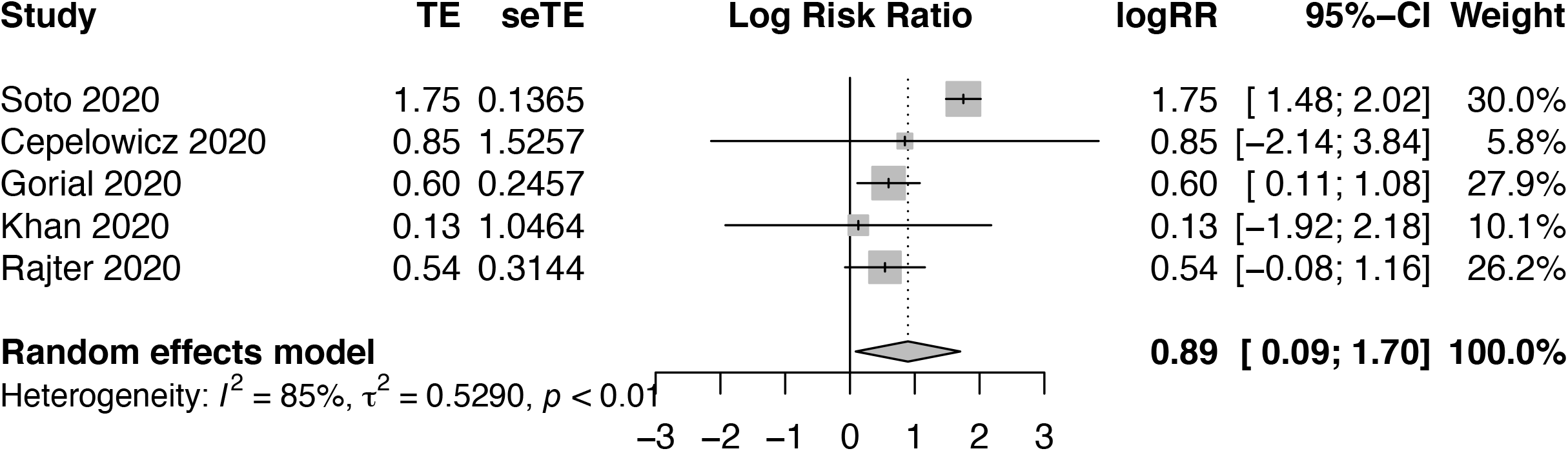
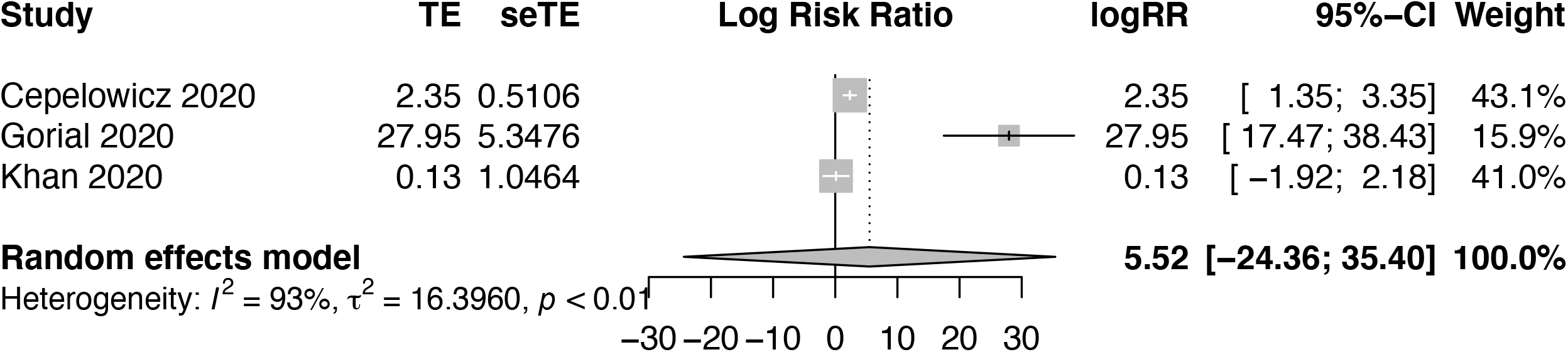
Forest plot for primary and secondary outcomes (a: Mortality; b: Recovered)

### Sensitivity analysis

No differences were found between the overall analysis and that proposed in the sensitivity analysis in terms of outcomes.

### Certainty of evidence in included studies

For certainty of evidence and evaluation of study quality, the GRADE recommendation was used. Two outcomes were assessed: Mortality (3607 participants, 5 retrospective studies), and recovery (397 participants, 3 pre-print retrospective studies). Both showed a very low certainty of the evidence, based on study design, risk of bias, inconsistency, indirectness and imprecision (Table 2).

**Table 2.**
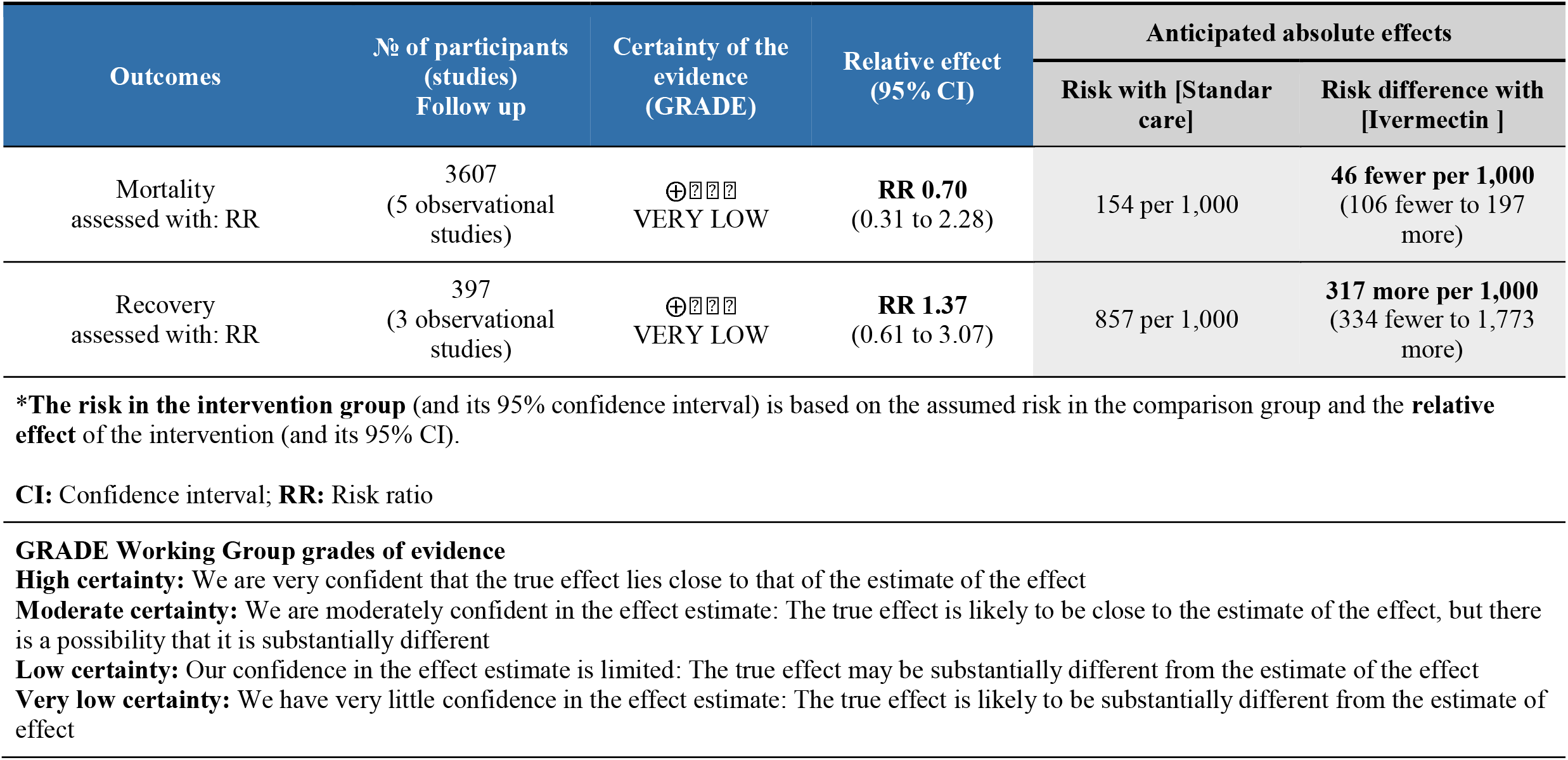
Summary of certainty evidence.

## Discussion

### Main results

We did not find a significantly reduction in the mortality and recovery of patients in the analyzed studies. It should be noted that the included studies are pre-print, so the information may vary and change the overall effect in our meta-analysis (although this trend may still be not significant).

Ivermectin has been widely used on the basis of having an antiviral effect against SARS-Cov-2 (27). In this regard, a study published in Australia showed in-vitro effectiveness of ivermectin in Vero-cells; however, its clinical application in humans is very doubtful (5). This study was rapidly adopted by clinical practice guidelines, recommending ivermectin for the treatment of patients hospitalized with COVID-19, especially in countries severely affected by the pandemic. For example, Peru, one of the countries hardest hit by the pandemic, included Ivermectin as a first-line treatment, even as prophylaxis (28).

The rationale to include this drug was based on its pharmacologic properties and application in other scenarios. Ivermectin belongs to the chemical group of avermectins, and is widely used in large animals for the treatment and control of parasitic infections, and to assist on the treatment of scabies and ticks. In humans, ivermectin has been used as a prophylactic drug in filariasis and as a therapeutic agent for scabies. It is a drug approved by the FDA and has shown to be safe in the recommended dosages (200 μg/kg) (29).

Despite the published theoretical information, there are not enough clinical trials to confirm the efficacy and safety of ivermectin for prophylaxis or treatment of patients with COVID-19. A systematic review was performed by Padhy et al (30), and analyzed the effects of ivermectin in 629 patients with COVID-19 (4 observational studies were included). The ivermectin-treated group had 233 mild cases and 104 moderate to severe cases. All-cause mortality was reduced in 2 out of the 3 included studies (OR 0.53, 95% CI 0.29-0.96). However, all these studies had a high risk of bias.

One of the limitations of the study conducted by Padhy et al. is that the overall effect of ivermectin on mortality was analyzed without considering the reported effect measure. Furthermore, the analysis was performed without transforming the individual effect (from OR to LogOR, for example), thus the OR reported was overestimated.

In another systematic review with networked meta-analysis, the effects of ivermectin on mortality were analyzed, and only two studies were included. The authors reported a very close statistical significance in terms of association of ivermectin with lower mortality (OR 0.15, 95% CI 0.04 to 0.57, p = 0.005); however, they pointed out that these data had very low certainty of evidence (31).

Despite the small amount of highly biased evidence that has been published supporting the efficacy of ivermectin, the specific human dose has not been established. Bray et al. evaluated in vitro whether an ivermectin concentration of 0.1 uM (instead of 5 uM) can inhibit SARS-Cov-2 (32). In clinical studies, the dose has ranged from 120 uM/kg to 200 uM/kg per dose in the intramuscular or oral form (33, 34). However, high doses for humans have not been approved (https://www.fda.gov/animal-veterinary/product-safety-information/faq-covid-19-and-ivermectin-intended-animals).

It is important to know that testing the efficacy of ivermectin in human clinical trials or observational study requires a previous evaluation in a dose-response trial, applying low dose (with less likelihood of pharmacological effect) and high dose relative to placebo. Given the lack of this type of studies, the ideal high dose of ivermectin has not been determined yet.

Our study has some limitations that are worthy to mention. First, regarding heterogeneity, we found that five out of the eight studies were done in inpatients, two studies focused on outpatients only, and one study focused on both outpatients and inpatients. Similarly, we have found differences between treatment arms, for example, five studies reported the use of ivermectin by itself compared to the standard of care, and the remaining studies used ivermectin in combination with other drugs (dexamethasone, hydroxychloroquine, azithromycin). Although the outcome in hospitalized patients was overall mortality or recovery time, in outpatients the outcome was appearance of symptoms of Covid-19, except in one study (Carvallo) that measured disease severity and mortality. As observed, there is clinical heterogeneity that makes it difficult to combine the estimates in a pooled estimate.

It is possible that the methodological heterogeneity and biases found in this systematic review and meta-analysis provided results inconsistent with reality. For this reason, it was proposed to meta-analyze the outcomes separately, assuming for each of them the model of random effects, and thus having greater precision in the effect. In spite of having statistical heterogeneity, the analysis of biases performed on the selected studies and the measurement of the size of the effect according to the outcome made a very close approximation to the reality.

Finally, the LogRR and confidence intervals for mortality and recovery were found to be non-significant, and by applying GRADE, we determined the certainty of the evidence for this estimated effect: The true effect is likely to be substantially different from the estimated effect. This systematic review and meta-analysis concludes that more randomized clinical trials need to be included in a meta-analysis, with fewer biases to approximate more to the real measurable effect. At the moment, there is no evidence that the use of ivermectin changes the clinical outcome of inpatients or outpatients.

## Data Availability

All database access data for the systematic search are available in our supplementary material.

## Notes

### Competing Interest Statement

The authors have declared no competing interest.

### Funding Statement

The authors declare that they have no conflicts of interest. This study has not been funded by any institution.

### Author Declarations

This study was a systematic review, so no ethics committee review was required for approval.

